# Longitudinal association of infant and early childhood body mass index with childhood and adolescent mental health: a Mendelian randomization study

**DOI:** 10.1101/2025.08.26.25334448

**Authors:** Xinyu Hu, Shan Luo, Chen Shen, Qian Yang

## Abstract

**Background:** Mental health issue during childhood and adolescence could have a lifelong influence on the quality of life, but its early-life risk factors are unclear. This study aims to explore the association of early childhood body mass index (BMI) on childhood and adolescent mental health.

**Methods:** We conducted a bi-directional two-sample Mendelian randomization (MR) study to investigate the association between infant and early childhood BMI and childhood and adolescence mental health disorders. Genetic instruments for BMI of 12 childhood age groups (from birth to 8 years old) were extracted from genome-wide association studies of up-to 28,681 European participants, and were used to proxy the primary childhood BMI exposures. Overall childhood BMI from an independent cohort was used as the validation exposure. Genetic associations with four childhood mental health disorders, including behavioural, emotional and social functioning disorders, aggression, and internalizing problems, were obtained from FinnGen and EAGLE consortia. The inverse-variance weighted or Wald ratio method was used as the discovery method, where MR-RAPS, dIVW, MR_cML were used as validation methods.

**Results:** In the primary analysis, tthe 1-year-old and 2-year-old BMI were robustly associated with behavioural and emotional disorders onset during childhood and adolescence (OR=1.10, 95%CI=1.01 to 1.19, P=0.0**24**; OR=1.12, 95%CI=1.002 to 1.24, P=0.046; respectively). These findings were replicated for emotional and social functioning disorders onset during childhood. BMI at 2 years old was robustly associated with aggression during childhood (OR=1.02, 95%CI=1.002 to 1.04, P=0.029). The analysis using independent childhood BMI data validated results for aggression. The bi-directional MR showed that none of the childhood mental health disorders had a reverse association with childhood BMI at any timepoint.

**Conclusions:** This study shows that BMI between 1-2 years old, and between 1.5-2 years old were robustly associated with behavioural and emotional disorder, and aggression, respectively. More attention is needed for early childhood weight control to prevent mental health disorders during childhood.

## Introduction

Globally, mental disorder during childhood and adolescence is a huge public health issue, influencing millions of children and adolescents. The Global Burden of Disease study showed that prevalences of at least one mental disorder differ across ages, ranging from 6.8% at 5-9 years to 14.0% at 15-19 years [1]. Compared to other non-communicable diseases, e.g. cardio-metabolic-kidney disease, the proportion of years lived with disability is much higher for mental disorders during childhood and adolescence [1].

Those mental disorders included several types of emotional and behavioural disorders (e.g., depression, anxiety, internalising and externalising difficulties) [2]. Several reasons were reported to cause childhood mental health disorders, including change of hormone levels during puberty, peer-pressure from school, comparison between family members [2]. In addition, brain is still in development during adolescence, making it sensitive to external stressors [3]. However, during to the complex interaction between genetics and environmental factors, there are many unknown risk factors for childhood mental health that are waiting to be identified.

The prevalence of overweight and obesity increases dramatically in children and adolescents around the world [4]. Many studies have showed adverse effects of increased childhood body mass index (BMI) and body size on increasing risks of type 2 diabetes, cardiovascular diseases and other chronic diseases [5–7]. In addition, adulthood BMI was reported to be associated with mental health such as depression [8]. Previously, a multi-country cross-sectional study of >1 million adolescents from Europe and North America observed a U-shaped association between adolescent BMI and mental health [9]. However, it is unclear whether BMI at an earlier life stage (from infancy to early childhood) is also associated with subsequent childhood and adolescent mental health issues.

Mendelian randomization (MR) is a statistical inference method that estimate the causal effect of an exposure on an outcome using genetic variants as instruments. This method provides an alternative approach to assess the association of childhood BMI with childhood and adolescent mental disorders. MR is less vulnerable to confounding than conventional observational study design, because genetic variants are randomly allocated at meiosis and cannot be influenced by the wide range of sociodemographic or other lifestyle factors [10]. In a two-sample MR setting, the genetic associations of the exposure and the outcome can come from independent studies within the same underlying population [11]. As a proof of principal, this approach has previously been used to evaluate causal associations of childhood BMI with psychiatric disorders during adulthood [12].

In this study, we aim to explore the association of infant and early childhood BMI with four childhood and adolescent mental disorders using two-sample MR (see **Figure 1**). Birth weight, childhood BMI at 10 different time points were use as exposures. The top MR findings were validated using an independent genome-wide association study (GWAS) of childhood BMI. Bidirectional MR was also conducted to minimise the possibility of reverse causality on the findings.

**Figure 1.**
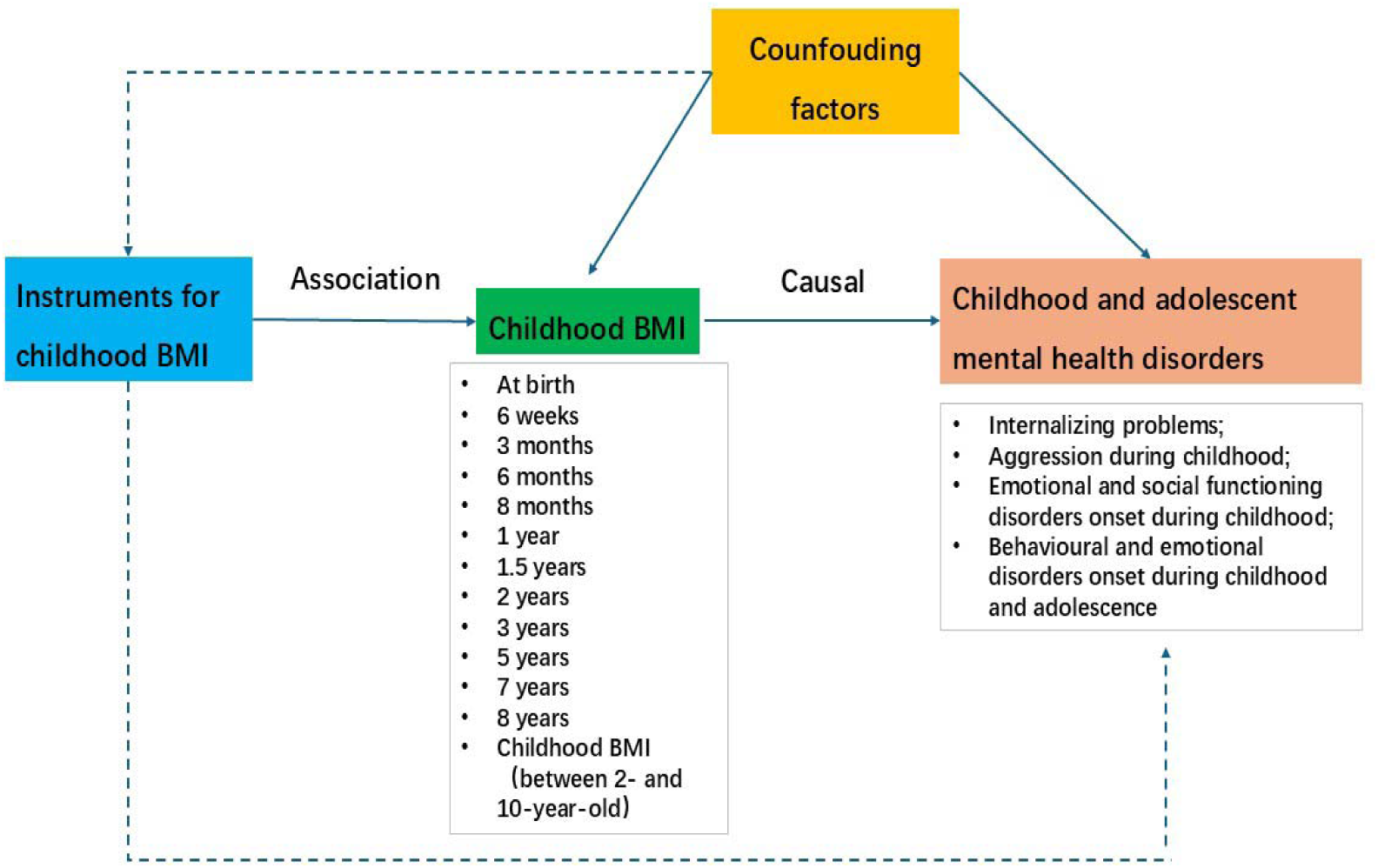
Conception and diagram of the study design. This Mendelian randomization study aims to explore the causal relationships between the association of early childhood body mass index (BMI) (exposure) on childhood and adolescent mental health (outcome). We included BMI of 12 childhood age groups (from birth to 8 years old) as exposure from 28,681 European participants, and 4 childhood mental health disorders, including behavioural, emotional and social functioning disorders, aggression and internalizing problems from 61,111 European children. Childhood BMI between 2- and 10-year-old from was used as an external validation exposure. The solid blue lines show effects that must exist and dashed blue lines represent effects that must not exist if an instrument variant is to be used to assess the causal effect of exposure on outcome.

## Methods

### Selection of infant and early childhood BMI exposures

To comprehensively explore the longitudinal influence of childhood BMI on childhood mental health, we selected 12 infant and early childhood BMI (i.e. body weight in kilograms divided by height or length in meters) as exposures of this study. This covered BMI from birth till 8 years old. Their genetic instruments for childhood BMI were obtained from a recent GWAS from the Norwegian Mother, Father and Child Cohort Study (MoBa), which included 28,681 children of European decent [13]. Childhood BMI at birth, 6 weeks, 3 months, 6 months, 8 months, 1 year, 1.5 years, 2 years, 3 years, 5 years, 7 years and 8 years were set as exposures separately. BMI was standardized to one standard deviation (SD) unit for all studies. In addition, childhood BMI between 2- and 10-year-old from Vogelezang et al. was used as an external validation exposure [14]. Its genetic instruments were obtained from the GWAS meta-analysis of 61,111 European children from Early Growth Genetics (EGG) consortium, including 552 MoBa participants.

### Selection of childhood mental health outcomes

To ensure the reliability of our findings, we obtained genetic associations with childhood and adolescent mental health from FinnGen release 12 [15] and EArly Genetics and Lifecourse Epidemiology (EAGLE) consortium [16]. Three childhood and adolescent mental health disorders were selected as main outcomes based on the data availability and sample size, including internalizing problems, aggression during childhood, as well as behavioural and emotional disorders onset during childhood and adolescence. The data with the largest available GWAS summary statistics of European ancestry were selected.

In more details, FinnGen is the nationwide network of Finnish biobanks linked to national electronic registries that provide information on diseases [15]. Behavioural and emotional disorders onset during childhood and adolescence is defined based on ICD-10 codes F90 to F95, including 9,640 cases and 490,708 controls (finn-b-R12_BEHEMOCHILD). Genetic associations with preschool internalizing problems were extracted from a GWAS meta-analysis of Benke et al. [16], including 4,596 children of European descent from three birth cohorts. Internalizing problems were consistently measured through Child Behavior Checklist (CBCL) and rated by their mothers. Genetic associations with aggression during childhood were obtained from a GWAS meta-analysis that included 87,485 children aged between 1.5 and 18 years from 29 studies [17]. Among these participants, 8,195 children were from MoBa. Aggression was measured through different questionnaires across studies, which were answered by parents, teachers, and/or participants themselves, but each study followed a standard operation protocol to ensure their GWAS summary statistics can be meta-analysed by the EAGLE consortium.

We also included emotional and social functioning disorders onset during childhood as a secondary outcome, which is defined based on ICD-10 codes F93 and F93. This outcome included 2,230 cases and 490,708 controls, and was used to further validate our results for behavioural and emotional disorders onset during childhood and adolescence.

### Statistical analyses

#### Mendelian randomization analyses

##### Instrument selection for infant and early childhood BMI

We applied a stringent instrument selection process to select valid instruments for our exposures. This process included the following steps: (1) ambiguous and palindromic variants were removed using the pre-defined approach included in the harmonization step of the TwoSampleMR R package [18]; (2) the genetic variants are associated with childhood BMI at a P value cut-off of 1×10^-5^; (3) the genetic variants were independently associated with childhood BMI, which passed a Linkage Disequilibrium (LD) clumping r^2^ threshold of 0.001; (4) the selected genetic instruments showed good instrument strength, which had F-statistics over 10.

##### Exploring associations of infant and early childhood BMI with childhood and adolescent mental health disorders

For the primary analysis, we conducted a two-sample MR analysis of 12 infant and early childhood BMI traits (i.e. at birth, 6 weeks, 3 months, 6 months, 8 months, 1 year, 1.5 years, 2 years, 3 years, 5 years, 7 years and 8 years) with four childhood and adolescent mental health disorders (emotional and social functioning disorders onset during childhood, behavioural and emotional disorders onset during childhood and adolescence, internalizing problems, as well as aggression during childhood) to detect the potential associations. We harmonized the summary datasets and outcomes through allele frequency alignment and strand orientation standardization. The MR-IVW or Wald ratio method was used as the discovery approach, given two or more instruments or one instrument, respectively [11]. Although the F-statistics of childhood BMI instruments were over 10, we still applied several sensitivity methods, including MR-RAPS [19], MR constrained maximum likelihood (cML) [20] and MR-debiased-IVW [21] approaches, to ensure robustness against potential biases arising from weak instrument effects. Bonferroni correction was used to adjust for multiple comparisons among four outcomes, giving a cutoff of P = 0.0125 (i.e. 0.05/4).

In addition to the primary analysis, we used childhood BMI between 2 and 10 years old as a validation exposure [14], and tested its association with the four childhood mental health disorders. Those associations also passed the validation analysis were treated as more reliable MR findings. The binary outcomes were demonstrated in odds ratio (OR) of mental health disorder per SD unit change of childhood BMI, while continuous outcomes were beta coefficient per SD unit change of childhood BMI.

##### Exploring bidirectional associations

We conducted bidirectional MR analyses of childhood and adolescent mental health disorders (as exposures) with infant and early childhood body weight (as outcomes) to minimise the possibility of reverse causality [22]. The instruments for childhood and adolescent mental health disorders were selected using the same four steps approaches as above. The MR-IVW was applied as the discovery approach, and MR-RAPS, MR cML and MR-debiased-IVW approaches were applied as sensitivity methods. We considered the associations of infant and early childhood BMI with childhood mental health disorders without reverse causal relationships as more reliable MR findings. The continuous outcomes were demonstrated in beta coefficient of infant and early childhood BMI per log odds change of binary traits or per SD unit change of mental health risk score.

##### Sensitivity analyses to test for MR assumptions

MR analysis relies on three key *assumptions*: relevance, exchangeability, and exclusion restriction [10].To ensure the genetic instruments fit the relevance assumption, we used the F-statistics to evaluate the validity of the selected instruments. Exposures with an F-statistic greater than 10 were considered to have sufficient strength to be included in the analysis [10]. For exchangeability assumption, the exposure and outcome data were both from European ancestry. For outcome data from FinnGen, we checked the difference in effect allele frequency (EAF) between the exposure and outcome GWAS. The absolute difference of EAF between exposure and outcome data < 0.1 was defined as minor departure of the population stratification. For exclusion restriction, we assessed horizontal pleiotropy using the MR-Egger regression [23], which applies weighted linear regression with an unconstrained intercept. The intercept signifies the mean pleiotropic effect and illustrates the average direct impact of a variant on the outcome. We also evaluated heterogeneity using Cochrane’s Q test to identify outlier variants [23].

MR analyses were conducted using R packages TwoSampleMR (version 0.6.6) in R (version 4.4.1) and mr.raps (version 0.2).

## Results

### Summary of instrument and outcome selection

For the 12 infant and early childhood BMI traits, one to 14 variants were selected as instruments to proxy those exposures (**Supplementary Table 1**). The F statistics for infant and early childhood BMI ranged from 34 to 59, suggesting sufficient instrument strength.

The four childhood and adolescent mental health disorders, including emotional and social functioning disorders onset during childhood, behavioural and emotional disorders onset during childhood and adolescence, internalizing problems, and aggression during childhood, were selected as outcomes in this study (**Supplementary Table 2**).

### Associations of infant and early childhood BMI with childhood and adolescent mental health disorders

For behavioural and emotional disorders onset during childhood and adolescence, per SD increased childhood BMI at 6 weeks old (OR=1.14, 95% confidence interval [CI]=1.02 to 1.28, P=0.024), at 1 year old (OR=1.10, 95%CI=1.01 to 1.19, P=0.024), and at 2 years old (OR=1.12, 95%CI=1.002 to 1.24, P=0.046) were both associated with this mental disorder (**Figure 2A**), though they cannot pass multiple comparison. These results were validated using MR-RAPS, dIVW and/or MR cML methods (**Supplementary Table 3**). In contrast, childhood BMI at other time points did not show sufficient evidence to support their associations with this outcome.

**Figure 2.**
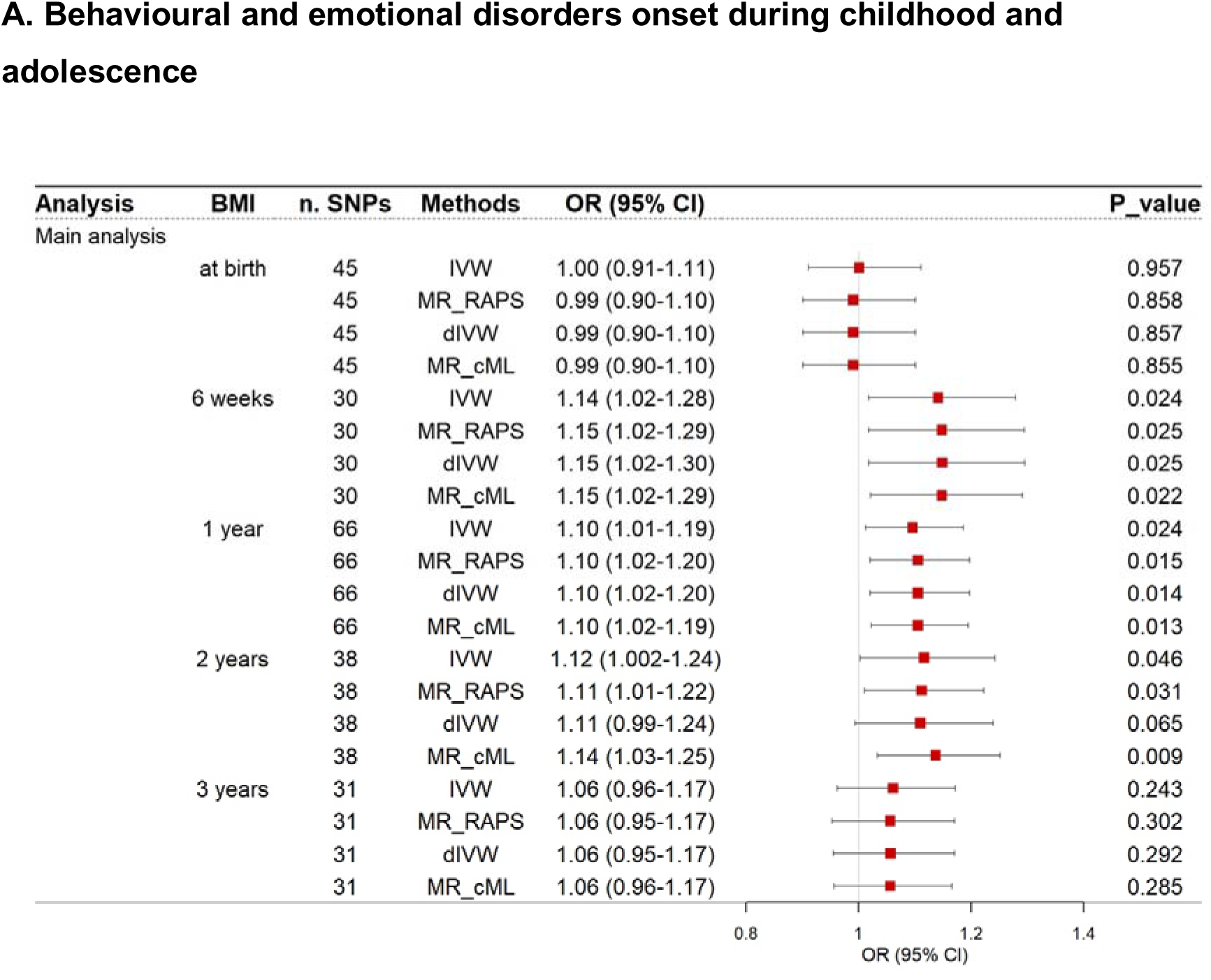
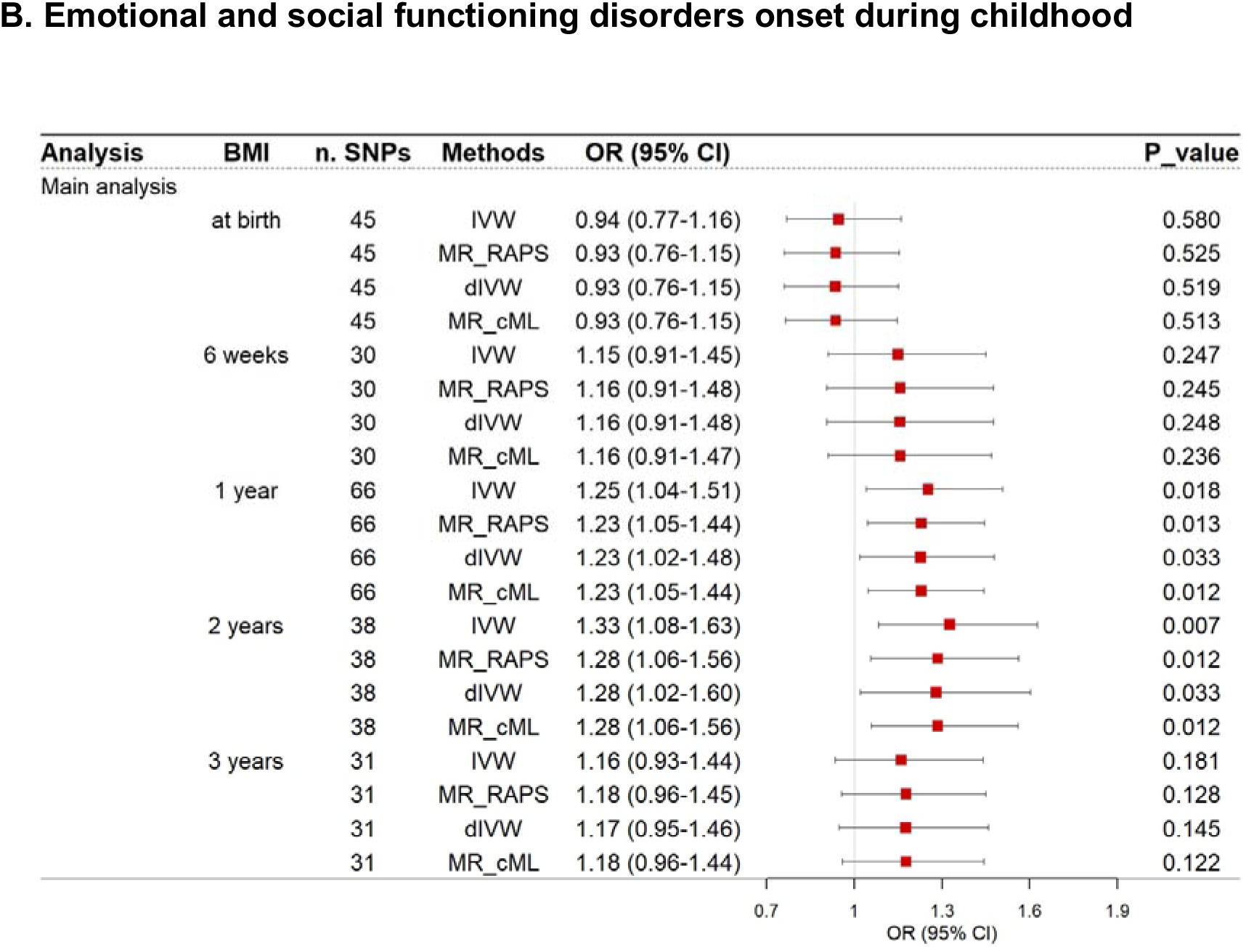
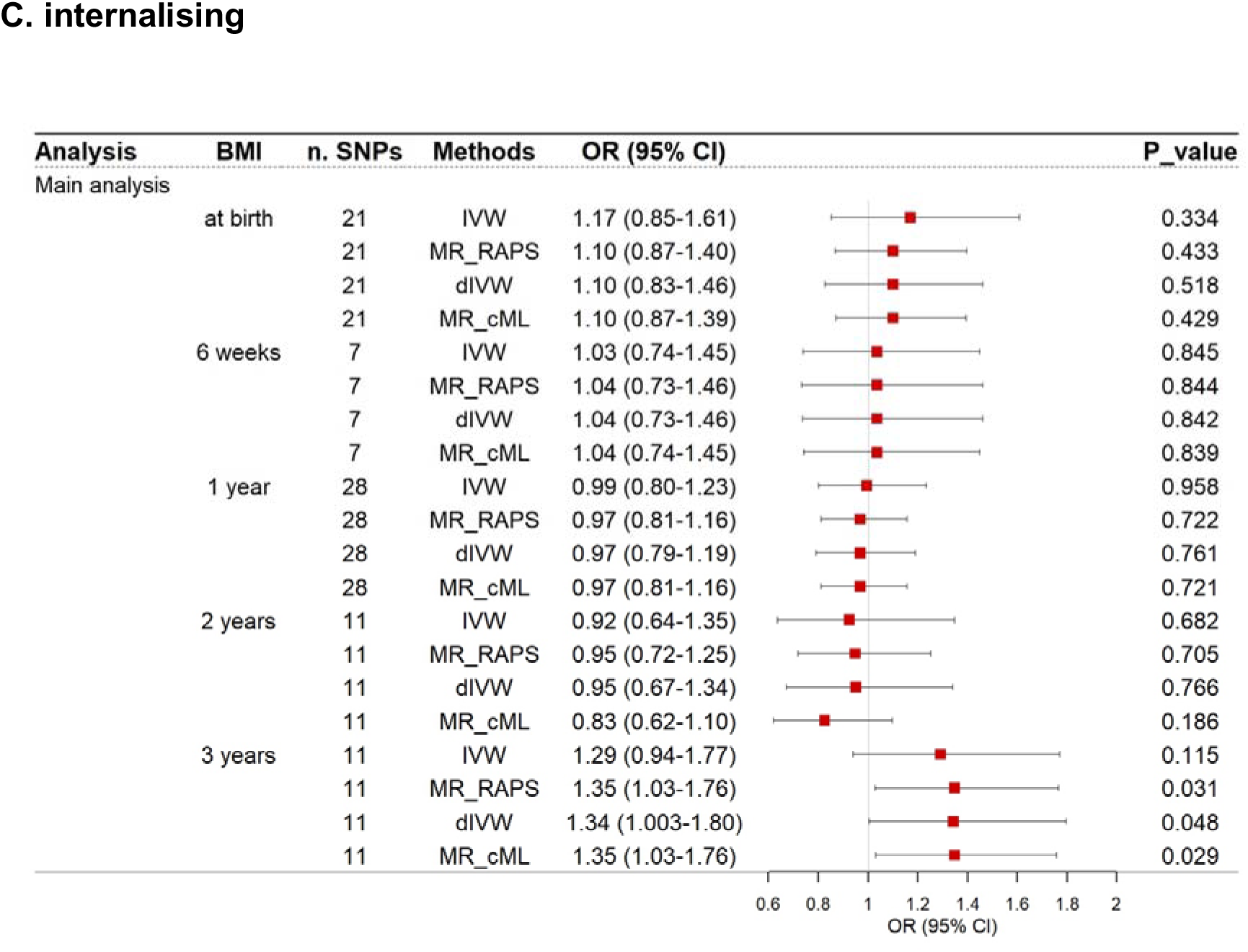
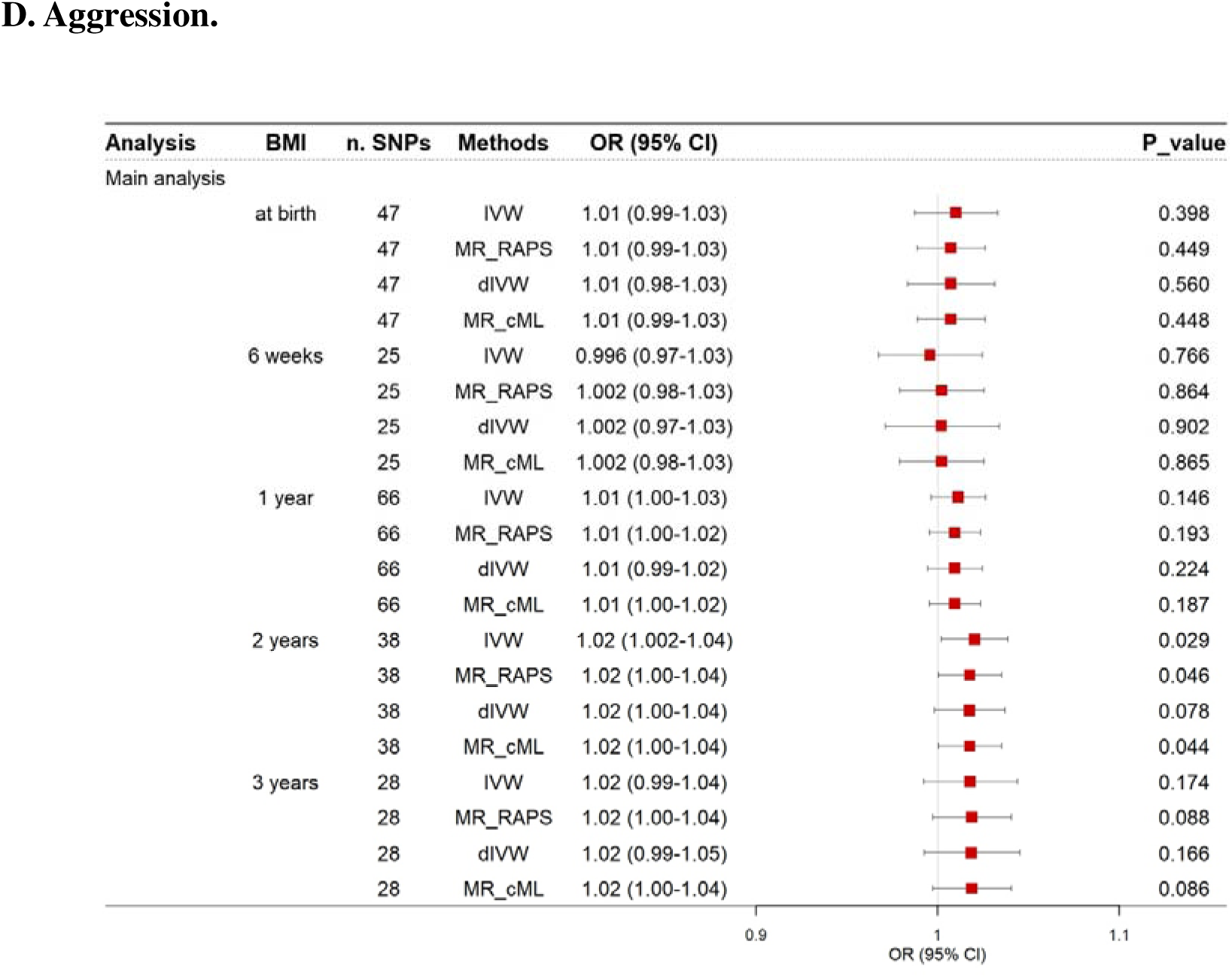
Causal associations of infant and early childhood BMI with childhood and adolescent mental health disorders. MR estimates were derived from the IVW method (the Wald ratio for those with only 1 IV variant) in the primary analysis and the MR_cML, the MR_RAPS, dIVW methods in the sensitivity analysis. The exposure IV SNPs in the main analysis were from the MoBa study including 28,681 children of European decent, and those in the validation analysis were from the EGG consortium incorporating 61,111 European children. The effect sizes for the difference stage of childhood BMI were associated with the four genetic predisposition to each of the four childhood and adolescent mental health disorders scaled to a SD, which was same as that in the previous genome-wide association study. The red squares represent ORs, and the error bars represent 95% CIs. Abbreviations: BMI, body mass index; SNP, single nucleotide polymorphism; CI, confidence interval; MR, mendelian randomization; cML, constrained maximum likelihood; RAPS, robust adjusted profile score; dIVW, Debiased Inverse-Variance Weighted; OR, odd ratio.

Using emotional and social functioning disorders onset during childhood as a validation, we generally observed similar association pattern as behavioural and emotional disorders despite weaker statistical power. We still observed consistently associations of BMI at 1 year old (OR=1.25, 95%CI=1.04 to 1.51, P=0.018) as well as association of BMI at 2 years old (OR=1.33, 95%CI=1.08 to 1.63, P=0.007) with this disorder (**Figure 2B**). In addition, BMI at 1.5 years old and 5 years old achieved marginal associations with disorder (**Supplementary Table 4**). By contrast, childhood BMI at other time points were not associated with emotional and social functioning disorders onset during childhood (**Supplementary Table 4**).

For internalizing problems, we observed that per SD unit increased BMI at 3 years old increased risks of internalizing problems by 29% (OR=1.29, 95%CI=0.94 to 1.77, P=0.115; **Figure 2C**) despite its 95%CI was overlapped with the null. This finding was validated using MR-RAPS, dIVW and MR cML methods (**Supplementary Table 5**). In contrast, childhood BMI at any other age groups did not show a significant association with internalizing problems.

For aggression during childhood, one SD unit increase in BMI at 2 years old (OR=1.02, 95%CI=1.002 to 1.04, P=0.029; **Figure 2D**) was significantly associated with aggression during childhood. These findings were validated using weighted median, MR-RAPS and MR cML methods (**Supplementary Table 6**), and the additional childhood BMI GWAS meta-analysis, which increased the robustness of the findings. In addition, this finding was also validated using the childhood BMI between 2- and 10-year-old in all the models (P values range from 0.0004 in MR_RAPS to 0.021 in MR_Egger; effect size is from 1.03-1.09). In contrast, insufficient evidence was found for associations of childhood BMI at other time points with this outcome.

### Associations of childhood and adolescent mental health disorders with infant and early childhood BMI

To verify the directionality of the associations, we conducted bidirectional MR for the above-mentioned significant associations. We identified 26, 40, 13, and 35 instruments for emotional and social functioning disorders onset during childhood, behavioural and emotional disorders onset during childhood and adolescence, internalizing problems, and aggression, respectively (**Supplementary Table 7A**).

The results showed that emotional and social functioning disorders onset during childhood was not associated with BMI at year 1 or year 3. Behavioural and emotional disorders onset during childhood and adolescence was not associated with BMI at year 1, year 3 or year 5. Internalizing problems were not associated with BMI at birth. Aggression during childhood was not associated with BMI at birth, year 3 or year 5. The above results were further validated using MR-Egger, weighted median, simple mode, weighted mode, MR_cML, MR-RAPS and dIVW (**Supplementary Table 7B**).

In summary, the bidirectional MR analysis showed that none of the childhood mental health disorders were associated with childhood BMI at any time point. This minimizes the possibility of the influence of reverse causality on our top MR findings.

### Falsification of Mendelian randomization assumptions

In this study, we report findings according to the STROBE-MR (Strengthening the Reporting of Mendelian Randomization Studies) guidelines [24] (**Supplementary Table 8**). To ensure the validity of our MR findings between childhood BMI and the tested mental health disorders, we falsified the following core MR assumptions:

For relevance assumption, we evaluated the strength of the instruments by checking the F-statistics. The F-statistics for all exposures were greater than 34.2 **(Supplementary Table 1, Supplementary Table 7A)**, indicating that the instruments are strongly associated with the exposures. MAR-RAPS, MR cML and dIVW methods were employed in addition to MR-IVW, which were designed to deal with weak instrument bias. We observed similar MR estimates among these methods, which again suggested that weak instrument bias is unlikely to be a key issue for our study.

For exchangeability assumption, the exposure and outcome GWAS were both from European ancestry. Although two outcome GWAS were from Finland, which have minor departure of population structure compared to the rest of the European population. By checking the allele frequency differences between our exposure and outcome GWAS, we observed no major differences (absolute difference of EAF between exposure and outcome data <0.1, **Supplementary Table 9**). This implies that population stratification is less likely to be an issue for our findings.

For exclusion restriction, we assessed the potential horizontal pleiotropy using MR-Egger regression. The intercept term from MR-Egger regression was not statistically different from zero (P ≥ 0.05). To increase the reliability of the findings, we applied random effect IVW model as the discovery method for all MR associations, which showed robust evidence to support the MR associations of childhood BMI on childhood mental health disorders (**Supplementary Table 3-6**).

## Discussion

A In this study, we estimated the associations of 12 infant and childhood BMI traits with four childhood and adolescent mental health disorders. Using large-scale genetic data of up to 28,681 European participants, we showed that one SD unit increase in BMI at 6 weeks old and at 1 years old was associated with an increased risk of behavioural and emotional disorders onset during childhood and adolescence by 14% and 10%, respectively, where the findings were replicated using extra exposure and outcome data. Increased BMI at 3 years old by one SD unit was associated with a 29% increased risk of internalizing problems, despite a wide 95%CI. In addition, one SD unit increase in BMI at 2 years old was significantly associated with an increased risk of aggression during childhood by 2%. Collectively, our results underline the importance of controlling BMI from infancy to early childhood to prevent those serious mental health disorders during childhood and adolescence. For behavioural and emotional disorders onset during childhood and adolescence, our results for the associations of higher BMI at 6-week-old and 1-year-old with higher risk of this outcome were directionally consistent with a systematic review and meta-analysis of 13 cross-sectional studies focusing on conduct disorder [25]. However, their findings were vulnerable to reverse causality and residual confounding, which could result in substantial heterogeneity in the meta-analysis. A recent large cross-sectional study focusing on obesity and attention-deficit hyperactivity disorders also supported our findings, though it could have the same sources of biases as this systematic review and meta-analysis [26]. We acknowledged that we combined all subtypes of behavioural and emotional disorders onset during childhood and adolescence together, while previous observational studies investigated these outcomes separately. For internalizing problems, our null associations found for childhood BMI after birth were largely consistent with a systematic review and meta-analysis of observational studies (N=65,608), in which neither overweight nor obesity was associated with childhood anxiety [27]. Similarly, null associations of overweight and obesity with depression, self-esteem, and body dissatisfaction were observed in the same systematic review and meta-analysis [27]. Inconsistent with our null associations, another systematic review and meta-analysis of observational studies showed an association of obesity with higher risk of depression in 42,913 children and adolescence [28]. However, considerate heterogeneities were observed in these meta-analyses, suggesting that their results need to be interpreted with caution. Our unfavourable association of BMI at birth with internalizing problems was less comparable with previous MR studies of birthweight on major depression in either childhood [29] or adulthood [30]. Both weight and BMI can be interpreted as proxies of fetal growth, and further studies with more accurate measures, e.g. via ultrasound, could help clarify its causal relationship with internalizing problems.

For aggression during childhood, our unfavourable associations were generally consistent with previous positive correlations between higher BMI and aggressive behaviours and externalizing problems observed in 8484 children [31]. Another observational study found no evidence to support an association of BMI at 7-10 years old (N=10,328) with aggressive behaviour [32], which was consistent with our null findings for BMI after 5 year old.

An old Chinese saying, echoed in William Wordsworth’s line “The child is father of the man”, suggests that a child’s personality is largely formed during early childhood and shapes the adulthood. Down this line, early interventions are needed to control risk factors for mental health disorders. Our findings have important public health implications by emphasising the need for development and testing of suitable and safe interventions to support young children maintaining a healthy BMI. This should be a key target to reduce the burden of mental health complications and improve the quality of life. More importantly, our study identified the window of intervention on childhood BMI, which could supplement evidence for clinical guidelines of both obesity and mental health during childhood. Our study has several strengths. First, our study systematically scanned associations of infant and childhood BMI at 12 time points, which comprehensively identified influences of early-stage obesity on later-stage mental health issues. Second, to inform future clinical guidelines and practices, we identified the optimal body weight intervention time points for childhood and adolescent mental health issues. By scanning the optimal body weight intervention time points, we successfully found that BMI before 3-year-old may maximise the risk reduction for mental health disorders during childhood and adolescence.

However, several limitations do exist. First, the sample size of age-specific childhood BMI was still limited. Therefore, we were not able to identify a huge number of genetic instruments to proxy childhood BMI at each time point. However, the instrument strength for childhood BMI at all time points is still exceeding an F-statistic of 10, which suggests our results are unlikely to be influenced by weak instrument bias [10]. In addition, we conducted a whole set of MR sensitivity methods such as debiased IVW and MR-RAPS, which were designed to deal with weak instrument bias. These methods suggested similar findings as MR-IVW results, which again validated the reliability of our instruments. Second, one mental health outcome, emotional and social functioning disorders onset during childhood, have a relatively limited number of cases, and therefore did not have enough power to identify significant MR findings as behavioural and emotional disorders. However, the pattern of MR estimates for the two outcomes is similar, which validated the reliability of our findings. Third, two of our outcome GWAS were from FinnGen study (e.g. behavioural and emotional disorders), which could suffer from minor department of population stratification [15], especially compared with the EGG GWAS meta-analysis of childhood BMI conducted across the Europe. By checking our data, we found that EAF of the childhood instruments were similar between the exposure (childhood BMI) and outcome (behavioural and emotional disorders) GWAS, with an absolute difference of EAF < 0.1 for all instruments. Therefore, our results were less likely to be influenced by population stratification.

Fourth, some two-sample MR results could be biased by sample overlaps due to inclusion of a few birth cohorts, e.g. Avon Longitudinal Study of Parents and Children and MoBa, in both BMI and EAGLE consortium’s GWAS [33]. Further studies are warranted to replicate our top findings. Fifth, we acknowledged potential measurement errors in our outcomes, which could have two sources. One could be misclassification of mental health disorders in FinnGen, especially when the percentage of emotional and social functioning disorders onset during childhood (0.5%) was lower than the 6.9% reported previously [34]. The other source could come from maternal rated CBCL for internalizing problems [17], in contrast to aggression rated by parents, teachers, and participants themselves. Sixth, we could not exam potential non-linear associations of BMI with the outcomes using MR due to the lack of individual participant data. In addition, recent debate on methodology of non-linear MR [35] suggested that conventional observational analyses were needed to answer such research question. Finally, our study was mainly conducted using data from European ancestry. Therefore, the generalisability across ancestries need to be tested in future studies.

Our results represent an assessment of associations of infant and early childhood BMI with childhood mental health disorders using MR. Our results provide evidence to support that children should control body weight at early stage of their life to prevent childhood mental health issues, including behavioural and emotional disorders, internalizing problems and aggression during childhood and adolescent. Our study may inform the development of early-stage intervention strategies to control childhood body weight, which will benefit both metabolic and mental health for later-stage adolescents.

## Supporting information

STROBE-MR

## Data Availability

Summary statistics from the Norwegian Mother, Father and Child Cohort Study, Early Growth Genetics consortium, FinnGen, and EArly Genetics and Lifecourse Epidemiology consortium are publicly available as declared in their publications.

## Contributors

QY conceived the research question, and designed the study. XYH analysed the data. XYH and QY wrote the first draft of the manuscript. QY supervised this study. All authors revised the manuscript critically for important intellectual content, and read and approved the final manuscript.

## Acknowledgements

The authors thank Norwegian Mother, Father and Child Cohort Study, Early Growth Genetics consortium, FinnGen, and EArly Genetics and Lifecourse Epidemiology consortium for sharing their summary-level data.

