## Supplementary material for "Longitudinal association of infant and early childhood body mass index with childhood and adolescent mental health: a Mendelian randomization study": STROBE-MR

**STROBE-MR checklist of recommended items to address in reports of Mendelian randomization studies**^1^ ^2^

| **Item No.** | **Section** | **Checklist item** | **Page No.** | **Relevant text from manuscript** |
| --- | --- | --- | --- | --- |
| 1 | **TITLE and ABSTRACT** | Indicate Mendelian randomization (MR) as the study’s design in the title and/or the abstract if that is a main purpose of the study | 1, 2 | Title: “Longitudinal associations of infant and early childhood body mass index with childhood and adolescent mental health: a Mendelian randomization study ”  Abstract: “**Methods**: We conducted a bi-directional two-sample Mendelian randomization (MR) study to investigate…” |
|  | **INTRODUCTION** |  |  |  |
| 2 | **Background** | Explain the scientific background and rationale for the reported study. What is the exposure? Is a potential causal relationship between exposure and outcome plausible? Justify why MR is a helpful method to address the study question | 3 | Rationale: paragraph 1, 2  Exposure: paragraph 3  Potential causal relationship: paragraph 3  Justify MR method: paragraph 4 |
| 3 | **Objectives** | State specific objectives clearly, including pre-specified causal hypotheses (if any). State that MR is a method that, under specific assumptions, intends to estimate causal effects | 4 | “In this study, we aim to explore the association of infant and early childhood BMI with four childhood and adolescent mental health disorders using two-sample MR (see **Figure 1**).” |
|  | **METHODS** |  |  |  |
| 4 | **Study design and data sources** | Present key elements of the study design early in the article. Consider including a table listing sources of data for all phases of the study. For each data source contributing to the analysis, describe the following: | 5 | “…we selected 12 infant and early childhood BMI (i.e. body weight divided by height or length) as exposures of this study.”  “…we obtained genetic associations with childhood and adolescent mental health from FinnGen release 12 [15] and EArly Genetics and Lifecourse Epidemiology (EAGLE) consortium [16]. Four childhood and adolescent mental health disorders were selected as main outcomes…”  We also provided Supplementary Table 2 to clearly listing sources of data. |
|  | a) | Setting: Describe the study design and the underlying population, if possible. Describe the setting, locations, and relevant dates, including periods of recruitment, exposure, follow-up, and data collection, when available. | 5 | “Their genetic instruments for childhood BMI were obtained from a recent GWAS from the Norwegian Mother, Father and Child Cohort Study (MoBa), which included 28,681 children of European decent [13]. Childhood BMI at birth, 6 weeks, 3 months, 6 months, 8 months, 1 year, 1.5 years, 2 years, 3 years, 5 years, 7 years and 8 years were set as exposures separately…”  “…including internalizing problems, aggression during childhood, emotional and social functioning disorders onset during childhood, as well as behavioural and emotional disorders onset during childhood and adolescence.” |
|  | b) | Participants: Give the eligibility criteria, and the sources and methods of selection of participants. Report the sample size, and whether any power or sample size calculations were carried out prior to the main analysis | 5 | “…. the Norwegian Mother, Father and Child Cohort Study (MoBa), which included 28,681 children of European decent [13].…”  “Emotional and social functioning disorders onset during childhood” is defined based on ICD-10 codes F93 and F94, which included 9,640 cases and 490,708 controls (finn-b-R12_EMOCHILD). “Behavioural and emotional disorders onset during childhood and adolescence” is defined based on ICD-10 codes F90 to F95, including 2,230 cases and 490,708 controls (finn-b-R12_BEHEMOCHILD). |
|  | c) | Describe measurement, quality control and selection of genetic variants | 6 | Please see the “Instrument selection for infant and early childhood BMI” part |
|  | d) | For each exposure, outcome, and other relevant variables, describe methods of assessment and diagnostic criteria for diseases | 5-6 | “…early childhood BMI (i.e. body weight divided by height or length) as exposures of this study.”  “Emotional and social functioning disorders onset during childhood” is defined based on ICD-10 codes F93 and F94,…. “Behavioural and emotional disorders onset during childhood and adolescence” is defined based on ICD-10 codes F90 to F95…). |
|  | e) | Provide details of ethics committee approval and participant informed consent, if relevant |  | Not relevant |
| 5 | **Assumptions** | Explicitly state the three core IV assumptions for the main analysis (relevance, independence and exclusion restriction) as well assumptions for any additional or sensitivity analysis | 7-8 |  |
| 6 | **Statistical methods: main analysis** | Describe statistical methods and statistics used | 6-8 |  |
|  | a) | Describe how quantitative variables were handled in the analyses (i.e., scale, units, model) | 7 | “The binary outcomes were demonstrated in odds ratio (OR) of mental health disorder per SD unit change of childhood BMI, while continuous outcomes were beta coefficient per SD unit change of childhood BMI. |
|  | b) | Describe how genetic variants were handled in the analyses and, if applicable, how their weights were selected | 7 |  |
|  | c) | Describe the MR estimator (e.g. two-stage least squares, Wald ratio) and related statistics. Detail the included covariates and, in case of two-sample MR, whether the same covariate set was used for adjustment in the two samples |  |  |
|  | d) | Explain how missing data were addressed |  | Not relevant |
|  | e) | If applicable, indicate how multiple testing was addressed | 7 | Bonferroni correction was used to adjust for multiple comparisons among four outcomes, giving a cutoff of P = 0.0125 (i.e. 0.05/4) |
| 7 | **Assessment of assumptions** | Describe any methods or prior knowledge used to assess the assumptions or justify their validity | 8 | Relevance: “Exposures with an F-statistic greater than 10 were considered to have sufficient strength to be included in the analysis”  “For exchangeability assumption, the exposure and outcome data were both from European ancestry.  “For exclusion restriction, we assessed horizontal pleiotropy using the MR-Egger regression [23], which applies weighted linear regression with an unconstrained intercept.”  “For exclusion restriction, we assessed horizontal pleiotropy using the MR-Egger regression [23],”  “We also evaluated heterogeneity using Cochrane’s Q test to identify outlier variants” |
| 8 | **Sensitivity analyses and additional analyses** | Describe any sensitivity analyses or additional analyses performed (e.g. comparison of effect estimates from different approaches, independent replication, bias analytic techniques, validation of instruments, simulations) | 7 | “The MR-IVW was applied as the discovery approach, and MR-RAPS, MR cML and MR-debiased-IVW approaches were applied as sensitivity methods.” |
| 9 | **Software and pre-registration** |  |  |  |
|  | a) | Name statistical software and package(s), including version and settings used | 8 | “MR analyses were conducted using R packages TwoSampleMR (version 0.6.6) in R (version 4.4.1) and mr.raps (version 0.2). ” |
|  | b) | State whether the study protocol and details were pre-registered (as well as when and where) |  | Not relevant |
|  | **RESULTS** |  |  |  |
| 10 | **Descriptive data** |  |  |  |
|  | a) | Report the numbers of individuals at each stage of included studies and reasons for exclusion. Consider use of a flow diagram |  | Not relevant |
|  | b) | Report summary statistics for phenotypic exposure(s), outcome(s), and other relevant variables (e.g. means, SDs, proportions) | 9 | “After selection, one to 14 variants were selected as instruments to proxy those exposures (**Supplementary Table 1**).  “The four childhood and adolescent mental health disorders, including emotional and social functioning disorders onset during childhood, behavioural and emotional disorders onset during childhood and adolescence, internalizing problems, and aggression during childhood, were selected as outcomes in this study (**Supplementary Table 2**).” |
|  | c) | If the data sources include meta-analyses of previous studies, provide the assessments of heterogeneity across these studies |  | Not relevant |
|  | d) | For two-sample MR:  i.  Provide justification of the similarity of the genetic variant-exposure associations between the exposure and outcome samples  ii.  Provide information on the number of individuals who overlap between the exposure and outcome studies | 11,5 | “By checking the allele frequency differences between our exposure and outcome GWAS, we observed no major differences (absolute difference of EAF between exposure and outcome data <0.1, Supplementary Table 9).”  “…Early Growth Genetics (EGG) consortium, including 552 MoBa participants.” |
| 11 | **Main results** |  |  |  |
|  | a) | Report the associations between genetic variant and exposure, and between genetic variant and outcome, preferably on an interpretable scale |  | Supplementary Table 1 |
|  | b) | Report MR estimates of the relationship between exposure and outcome, and the measures of uncertainty from the MR analysis, on an interpretable scale, such as odds ratio or relative risk per SD difference |  | “We still observed consistently marginal associations of BMI at 1 year old (OR=1.43, 95% confidence interval [CI]=1.05 to 1.94, P=0.03; Figure 2) with this disorder using both MR-RAPS and MR_cML methods; as well as association of BMI at 3 years old (OR=1.97, 95%CI=1.00 to 3.87, P=0.05; Figure 2) using MR-IVW method.” |
|  | c) | If relevant, consider translating estimates of relative risk into absolute risk for a meaningful time period |  | Not relevant |
|  | d) | Consider plots to visualize results (e.g. forest plot, scatterplot of associations between genetic variants and outcome versus between genetic variants and exposure) |  | Figure 2 |
| 12 | **Assessment of assumptions** |  |  |  |
|  | a) | Report the assessment of the validity of the assumptions | 11 | Paragraph1-4 for “Test for Mendelian randomization assumptions” |
|  | b) | Report any additional statistics (e.g., assessments of heterogeneity across genetic variants, such as *I^2^*, Q statistic or E-value) | 11 | “The F-statistics for all exposures were greater than 34.2 **(Supplementary Table 1, Supplementary Table 7A)**,”  “The intercept term from MR-Egger regression was not statistically different from zero (P ≥ 0.05).” |
| 13 | **Sensitivity analyses and additional analyses** |  |  |  |
|  | a) | Report any sensitivity analyses to assess the robustness of the main results to violations of the assumptions | 11 | “MAR-RAPS, MR cML and dIVW methods were employed in addition to MR-IVW, which were designed to deal with weak instrument bias.” |
|  | b) | Report results from other sensitivity analyses or additional analyses |  |  |
|  | c) | Report any assessment of direction of causal relationship (e.g., bidirectional MR) | 10 | bidirectional MR |
|  | d) | When relevant, report and compare with estimates from non-MR analyses |  | Not relevant |
|  | e) | Consider additional plots to visualize results (e.g., leave-one-out analyses) |  | Figure 2 |
|  | **DISCUSSION** |  |  |  |
| 14 | **Key results** | Summarize key results with reference to study objectives | 12 | The first paragraph in Discussion part |
| 15 | **Limitations** | Discuss limitations of the study, taking into account the validity of the IV assumptions, other sources of potential bias, and imprecision. Discuss both direction and magnitude of any potential bias and any efforts to address them | 13,14 | “First, the sample size of age-specific childhood BMI was still limited.”  “Second, one mental health outcome, emotional and social functioning disorders onset during childhood, have a relatively limited number of cases, and therefore did not have enough power to identify significant MR findings…”  “Third, two of our outcome GWAS were from FinnGen study (e.g. behavioural and emotional disorders), which could suffer from minor department of population stratification…”  “Fourth, some two-sample MR results could be biased by sample overlaps due to inclusion of a few birth cohorts,…”  “Fifth, we acknowledged potential measurement errors in our outcomes, which could have two sources.”  “Finally, our study was mainly conducted using data from European ancestry.” |
| 16 | **Interpretation** |  |  |  |
|  | a) | Meaning: Give a cautious overall interpretation of results in the context of their limitations and in comparison with other studies | 12-13 | The second paragraph. “our results for the associations of higher BMI at 1- and 3-year-old with higher risk of this outcome were directionally consistent with a systematic review and meta-analysis of 13 cross-sectional studies focusing on conduct disorder [25]. …” |
|  | b) | Mechanism: Discuss underlying biological mechanisms that could drive a potential causal relationship between the investigated exposure and the outcome, and whether the gene-environment equivalence assumption is reasonable. Use causal language carefully, clarifying that IV estimates may provide causal effects only under certain assumptions | 13 | “Both weight and BMI can be interpreted as proxies of fetal growth, and further studies with more accurate measures, e.g. via ultrasound, could help clarify its causal relationship with internalizing problems.” |
|  | c) | Clinical relevance: Discuss whether the results have clinical or public policy relevance, and to what extent they inform effect sizes of possible interventions | 13 | “This should be a key target to reduce the burden of mental health complications and improve the quality of life. More importantly, our study identified the window of intervention on childhood BMI, which could supplement evidence for clinical guidelines of both obesity and mental health during childhood.” |
| 17 | **Generalizability** | Discuss the generalizability of the study results (a) to other populations, (b) across other exposure periods/timings, and (c) across other levels of exposure | 14 | “Finally, our study was mainly conducted using data from European ancestry. Therefore, the generalisability across ancestries need to be tested in future studies.” |
|  | **OTHER INFORMATION** |  |  |  |
| 18 | **Funding** | Describe sources of funding and the role of funders in the present study and, if applicable, sources of funding for the databases and original study or studies on which the present study is based | 15 | “This work was not supported by any specific grant. QY is an honorary member of a unit funded by UK Medical Research Council (MC_UU_00032/5)” |
| 19 | **Data and data sharing** | Provide the data used to perform all analyses or report where and how the data can be accessed, and reference these sources in the article. Provide the statistical code needed to reproduce the results in the article, or report whether the code is publicly accessible and if so, where | 15 | “Summary statistics from the Norwegian Mother, Father and Child Cohort Study, Early Growth Genetics consortium, FinnGen, and EArly Genetics and Lifecourse Epidemiology consortium are publicly available as declared in their publications. All scripts are available via GitHub ….” |
| 20 | **Conflicts of Interest** | All authors should declare all potential conflicts of interest | 15 | “None” |

This checklist is copyrighted by the Equator Network under the Creative Commons Attribution 3.0 Unported (CC BY 3.0) license.

1.	Skrivankova VW, Richmond RC, Woolf BAR, Yarmolinsky J, Davies NM, Swanson SA, et al. Strengthening the Reporting of Observational Studies in Epidemiology using Mendelian Randomization (STROBE-MR) Statement. JAMA. 2021;under review.

2.	Skrivankova VW, Richmond RC, Woolf BAR, Davies NM, Swanson SA, VanderWeele TJ, et al. Strengthening the Reporting of Observational Studies in Epidemiology using Mendelian Randomisation (STROBE-MR): Explanation and Elaboration. BMJ. 2021;375:n2233.
